# COVID-19 outbreaks in Australia during a period of high epidemic control, 2020

**DOI:** 10.1101/2022.02.07.22270575

**Authors:** Freya G Hogarth, Rachel Nye, Nevada Pingault, Simon Crouch, Cushla Coffey, Kylie Smith, Rowena Boyd, Catherine Kelaher, Tracie Rentin, Moira C Hewitt, Ben Polkinghorne, Martyn D Kirk

**Affiliations:** Australian Government Department of Health, Canberra, Australian Capital Territory, Australia; National Centre for Epidemiology and Population Health, The Australian National University, Canberra, Australian Capital Territory, Australia; Western Australian Department of Health, Perth, Western Australia, Australia; Victorian Department of Health, Melbourne, Victoria, Australia; Queensland Health, Brisbane, Queensland, Australia; Tasmanian Department of Health, Hobart, Tasmania, Australia; NT Department of Health, Darwin, Australia; New South Wales Department of Health, Sydney, Australia

## Abstract

To describe characteristics of COVID-19 outbreaks in Australia to guide policy development for mitigation of future outbreaks, we conducted a retrospective analysis of COVID-19 outbreaks affecting two or more people reported to COVID-Net—an Australian national surveillance network—from 28 January until 27 December 2020. The COVID-Net surveillance network covered all Australian states and territories, with an estimated population of 25,649,985 persons as at 31 June 2020. We reported the epidemiology of COVID-19 outbreaks in Australia, including the setting in which they occurred, size, and duration.

853 outbreaks of COVID-19 were reported; associated with 13,957 confirmed cases, of whom 2,047 were hospitalised, and 800 died. The pattern of outbreaks followed a similar trend to the epidemic in Australia, defined by two distinct peaks in mid-March and July. Victoria reported the greatest number of outbreaks across all settings aligned with the second wave of infections. Outbreaks most commonly occurred in the workplace/industry setting (22%, 190/853), followed by education (14%, 122/853), residential aged care (13%, 114/853) and hospitals (10%, 83/853). The majority (40%, 340/853) of outbreaks had 6 to 24 cases, and the median outbreak duration increased in proportion with the number of associated cases.

This report summarising COVID-19 outbreaks in Australia identifies settings of highest risk. Surveillance of outbreaks informs our understanding of transmission dynamics in Australia relative to national and jurisdictional interventions. For settings that are high risk for COVID-19, it is important to prioritise planning, surveillance, and implementation of control measures.

## Introduction

Severe acute respiratory syndrome coronavirus 2 (SARS-CoV-2), the virus that causes coronavirus disease 2019 (COVID-19), has spread rapidly around the world since it was first identified in December 2019 [1]. Basic reproductive number (R0) estimates for SARS-CoV-2 range from 2–4; similar to or higher, than for severe acute respiratory syndrome and influenza virus [2, 3]. Because SARS-CoV-2 is highly infectious, COVID-19 outbreaks occur when people come into close contact with cases of COVID-19 [2, 4]. Early reports indicated that SARS-CoV-2 predominantly spread via respiratory droplets and fomites, emerging literature suggests transmission via aerosols is important [2, 5].

Outbreak investigations provide insights into SARS-CoV-2 transmission dynamics, including the risk in different settings and as an indicator of community transmission. Indoor and enclosed spaces where persons are in close proximity and there is low airflow increase the risk of SARS-CoV-2 infection [6, 7]. In these environments, behaviours such as sneezing, coughing, shouting, and singing result in greater volume of droplets suspended for longer periods [2, 8]. These environments may result in superspreader events, where one infectious individual infects a large number of secondary cases [8, 9]. Superspreader events are difficult to predict and result in epidemic growth that challenge control efforts [6, 10, 11].

In Australia during March 2020, the first epidemic wave of COVID-19 was characterised by cases arriving on international cruise ships and by air. This was followed by local transmission in settings where people congregate. Australian state and territory governments instituted extensive non-pharmaceutical interventions, along with case isolation, and tracing and quarantine of contacts. The Communicable Diseases Network Australia (CDNA) developed a national surveillance plan that included capturing data on outbreaks [12].

The Australian Government established COVID-Net, a surveillance network to support jurisdictional investigation of COVID-19 outbreaks and collect national surveillance data. States and territories have reported outbreaks and clusters of COVID-19, with Victoria experiencing a second wave due to community transmission.

In this report, we characterise outbreaks using surveillance data during a setting of high epidemic control in Australia. We examine where outbreaks occurred and identify factors associated with transmission.

## Methods

### COVID-19 outbreak definition

We defined a case of COVID-19 as a person testing positive to a SARS-CoV-2 real-time reverse transcription-polymerase chain reaction (RT-PCR) assay, and a primary case as the first known case of the outbreak [2]. If a case in one outbreak became the primary case in a secondary outbreak, the case may have been included in both outbreaks.

We defined a COVID-19 outbreak as two or more cases (who did not reside in the same household) among a specific group of people and/or over a specific period of time where illness was associated with a common source of infection. We defined a multi-jurisdictional outbreak as involvement of two or more jurisdictions in the outbreak, regardless of where exposure occurred.

### Data sources

Our report includes outbreak data reported by all jurisdictions to COVID-Net, from 28 January until 27 December 2020 inclusive, while New South Wales (NSW) reported outbreaks from 5 July 2020 onwards. Local public health units investigated outbreaks and collected data that were reported by COVID-Net epidemiologists using a REDCap (Research Electronic Data Capture) database hosted at the Australian Government Department of Health [13].

Data collected for each outbreak included place of acquisition of the primary case, and number of cases, hospitalisations, and deaths. We categorised outbreaks as one of 13 settings and 56 sub-settings where the outbreak primarily occurred (S2 Appendix). The outbreak duration was derived by the number of days between the first and last positive specimen collected in the outbreak, plus 14 days from the last specimen date.

We obtained the total number of COVID-19 cases notified up to 27 December 2020 from the National Notifiable Disease System (NNDSS).

### Data analysis

We performed analyses in R 4.0.0 on a dataset extracted on 28 January 2021. We used population data from the Australian Bureau of Statistics Estimated Resident Population (as at 31 June 2020) to estimate rates of outbreaks by jurisdiction.

We analysed outbreak data by two distinct peaks relative to the 1 June 2020: from 28 January 2020 to 31 May 2020, and 1 June 2020 to 27 December 2020. We refer to these two periods as the first wave and second wave, respectively.

### Ethics

Surveillance activities were conducted for and on behalf of the Australian Government Department of Health under the National Health Security Act 2007. The activities were approved by the Australian National University Human Ethics Committee [HREC/17/ANU/909].

## Results

### National overview

COVID-Net sites reported 853 outbreaks of COVID-19, associated with 13,957 confirmed cases, of whom 2,047 were hospitalised, and 800 died. The median number of cases associated with these outbreaks was 6 (range 2–331). As at 27 December, 49% (13,957/28,312) of all confirmed COVID-19 cases notified in Australia, excluding NSW outbreak cases prior to 5 July 2020, were associated with an outbreak affecting ≥2 persons.

The overall rate of COVID-19 outbreaks during the surveillance period was 3.3 outbreaks per 100,000 population. The highest rates were in Victoria (10.3 per 100,000 population; Table. 1). Victoria reported the largest proportion of outbreaks (81%, 691/853) and associated cases (83%, 11,610/13,957), followed by NSW, excluding cases prior to 5 July 2020 (6%, 54/853 outbreaks, 5%, 642/13,957 cases). Overall, multi-jurisdictional outbreaks had the second highest proportion of cases (6%, 875/13,957), with a median number of cases of 15 (range 2 – 331; Table 1).

**Table 1.** Outbreaks of COVID-19 by state and territory, Australia, 28 January – 27 December 2020.

| State or territory | Number of outbreaks | Median size (persons) per outbreak | Number hospitalised | Outbreak rate per 100 000 population |
| --- | --- | --- | --- | --- |
| ACT | 2 | 5 | 1 | 0.5 |
| NSW * | 54 | 9 | N/A | 0.7 |
| NT | 0 | - | - | - |
| Qld | 36 | 3 | 78 | 0.7 |
| SA | 23 | 4 | 26 | 1.3 |
| Tas | 4 | 5 | 30 | 0.7 |
| Vic | 691 | 6 | 1,660 | 10.3 |
| WA | 21 | 5 | 76 | 0.8 |
| MJO | 22 | 15 | 176 | - |
| Total | 853 | - | 2,047 | - |

\* NSW commenced reporting COVID-19 outbreaks from 5 July 2020 onwards. COVID-19: coronavirus disease 2019; ACT: Australian Capital Territory; NSW: New South Wales; NT: Northern Territory; Qld: Queensland; SA: South Australia; Tas: Tasmania; Vic: Victoria; WA: Western Australia, MJO: Multi-jurisdictional outbreak.

### National trend

The pattern of outbreaks followed the wider epidemic in Australia, defined by two distinct waves relative to 1 June 2020 (Fig 1). Outbreaks in the first wave peaked in March 2020 and subsided by 31 May 2020, at which time COVID-Net sites reported 133 outbreaks. In the second wave, the source of infection for the primary case was locally acquired in 98% (705/720) of outbreaks compared to only 50% (67/133) in the first wave (odds ratio 47.7, 95% confidence interval 25.8–88.2), reflecting the impact of international travel restrictions progressively introduced from 15 March 2020.

**Fig 1.**
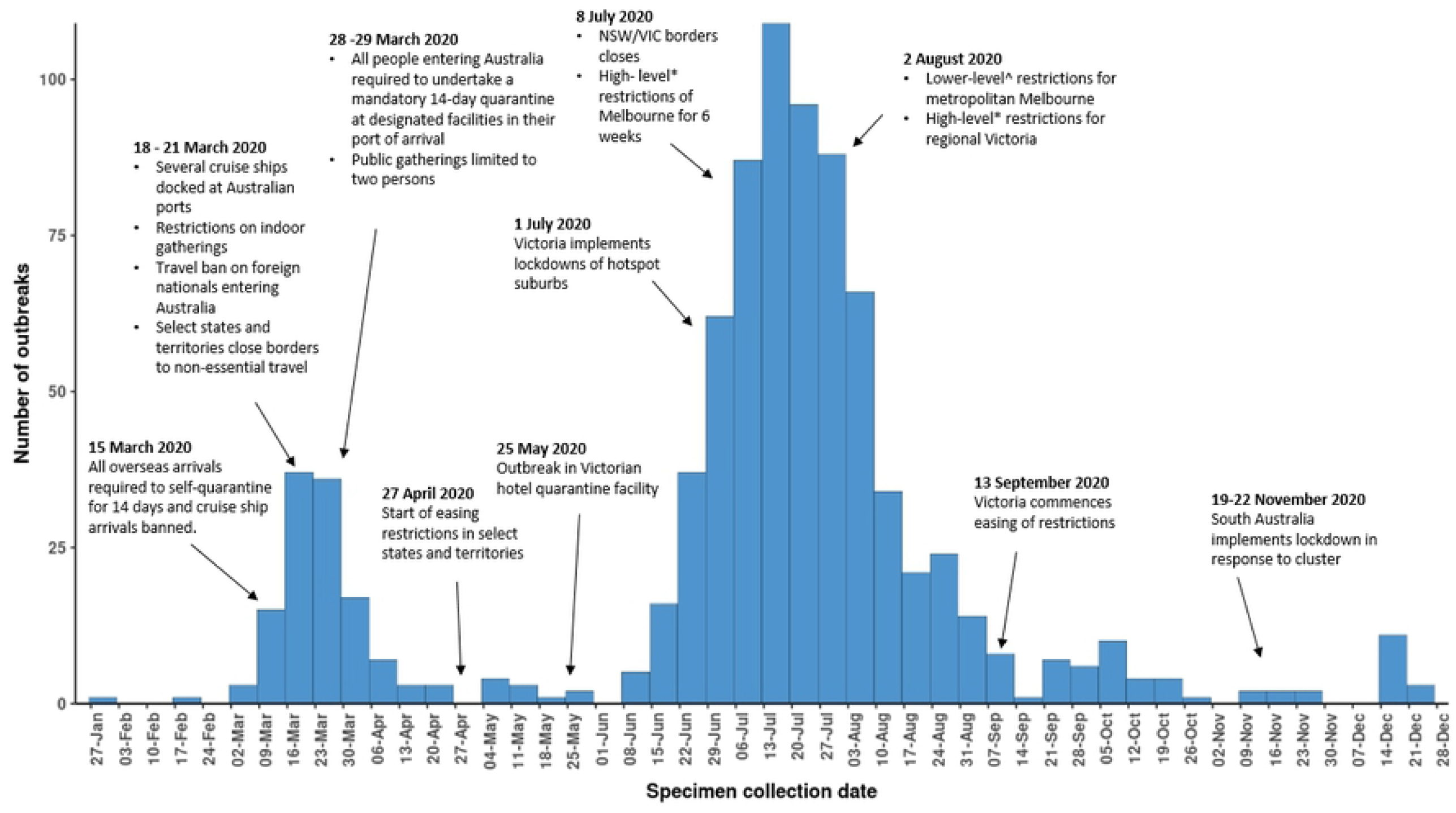
Number of outbreaks by specimen collection date of the primary case in the outbreak, Australia, 28 January – 27 December 2020, with timing of key public health measures. Adapted from “COVID-19 notifications in Australia: Epidemiology Report 30” by COVID-19 National Incident Room Surveillance Team, 2020, Comm Dis Intel, 44, https://doi.org/10.33321/cdi.2020.44.91. Copyright 2020 by Commonwealth of Australia * High-level restrictions include: compulsory mask-wearing in public; restrictions to travel within 5km from home; night-time curfew all non-essential retail and services closed; restaurants and cafes takeaway only; work from home unless essential worker; and online schooling. ^Lower-level restriction include: compulsory mask-wearing in public; exercise for two hours per day with one other person; restrictions to travel within 5km from home; some non-essential retail open; work from home unless essential worker; schools reopen for vulnerable children or whose parents cannot work from home.

The number of outbreaks in the second wave was much larger, characterised by sustained community transmission in one state - Victoria. The number of outbreaks reported each week increased exponentially (Fig 1). The peak in the number of COVID-19 cases was observed nationally in August 2020 when outbreak numbers had started to decline.

### Outbreak size and duration

The majority (40%, 340/853) of outbreaks had between 6 and 24 cases, and many (32%, 275/853) were smaller outbreaks of 3 to 5 cases. The median outbreak duration increased proportionally to the number of cases associated with an outbreak (Table 2).

**Table 2.** Outbreak size by duration, Australia, 28 January – 27 December 2020.

| Size<br>(number of cases) | Number of outbreaks<br>(%) | Median duration<br>(days) | Duration range<br>(min – max) |
| --- | --- | --- | --- |
| 2 | 122 (14%) | 18 | 14 – 86 |
| 3 to 5 | 275 (32%) | 21 | 14 – 113 |
| 6 to 24 | 340 (40%) | 29 | 15 – 132 |
| 25 to 49 | 56 (7%) | 38 | 14 – 99 |
| 50 to 74 | 16(2%) | 51 | 27 – 102 |
| 75 to 99 | 15 (2%) | 54 | 33 – 111 |
| 100+ | 29 (3%) | 64 | 21 – 138 |

The largest multi-jurisdictional outbreak occurred on a cruise ship among passengers and crew (n=854)[14]. Of cases, 296 passengers were reported by COVID-Net (excluding NSW), while 367 NSW passenger cases and 197 crew cases were documented in the Special Commission of Inquiry into the Ruby Princess [10]. The number of secondary cases among Australian residents who were infected by passengers and crew on board the ship was difficult to ascertain for this outbreak. The largest single jurisdictional outbreak occurred in a residential aged care facility (RACF) (n=301) and lasted for 86 days.

### Outbreak setting

During the two study periods, there were four settings where 60% (509/853) of COVID-19 outbreaks and 67% (9,350/13,957) of outbreak-associated cases occurred (S1 Appendix; S2 Appendix). Workplace/industry settings comprised 22% (190/853) of outbreaks and 11% (1,560/13,957) of cases (median=5, range 2–69). These outbreaks most frequently occurred in offices or call centres (n=31), unspecified workplaces (n=28), and non-food manufacturing (n=26). Educational settings comprised 14% (122/853) of outbreaks and 9% (1,189/13,957) of cases (median=6, range 2 – 209). Outbreaks in education settings occurred in secondary schools (n=52), primary schools (n=31), and combined primary-secondary schools (n=18). Outbreaks in RACF comprised 13% (114/853) of all outbreaks and 38% (5,275/13,957) of cases (median=11, range 2 – 301). Sixty-nine per cent (79/114) of these RACF outbreaks occurred between 1 June and 31 July 2020. Lastly, 10% (83/853) of outbreaks occurred in hospitals affecting 10% (1,326/13,957) of outbreak-associated cases. Among these, outbreaks most frequently occurred in acute care hospitals (n=69).

In the first wave, 33% (44/133) of COVID-19 outbreaks were reported in travel and transport settings, particularly among travel groups (n=21) and on cruise ships (n=14) (Fig 2). Locally acquired cases associated with these outbreaks occurred across the country. Of the 22 multi-jurisdictional outbreaks, 9 were associated with an outbreak on a cruise ship. Outbreaks among family and social gatherings (27 outbreaks; median=8, range 2–45) were prominent during the early stages of the epidemic when non-pharmaceutical interventions were not introduced. In the second wave, outbreaks and associated cases were predominantly reported from Victoria and were reported from a variety of settings (Fig 2). Notably, there was a substantial increase in the number of outbreaks in workplace/industry and educational settings compared to the first wave (S1 Appendix).

**Fig 2.**
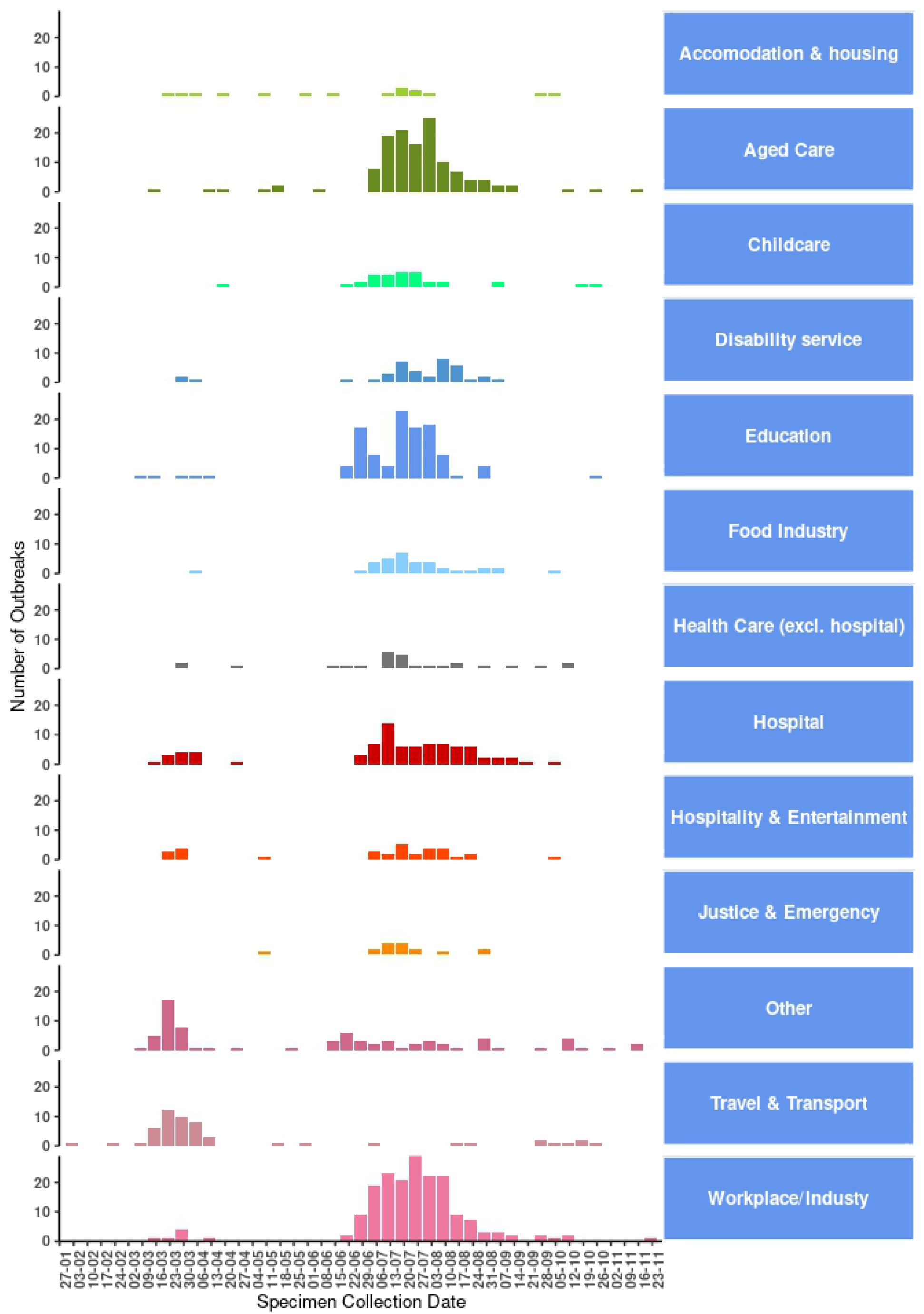
Number of COVID-19 outbreaks, by setting and specimen collection date for the index case per outbreak, Australia, 27 December 2020.

## Discussion

Australia exerted a high degree of COVID-19 control early in the pandemic through extensive non-pharmaceutical interventions. Outbreaks alerted health authorities to settings that required specific restrictions, such as international travel and large social gatherings. Outbreak data in the second wave strongly reflects the situation in Victoria. Overall, 81% (691/853) of outbreaks occurred in Victoria, and resulted in additional important transmission settings that were strongly influenced by a change in demographic characteristics of cases in the second wave.

In the first wave, approximately 50% (66/133) of outbreaks were linked to the introduction of infection from someone who travelled to Australia from overseas — including those at sea. SARS-CoV-2 is easily transmitted on board cruise ships due to high population density, crowded communal and living areas, and shared sanitary facilities [15]. Initially, infected cruise ships passengers disembarked at a range of ports around Australia—in many instances continuing travel across the country whilst infectious. A coordinated response by state and territory health authorities was required to rapidly detect and investigate COVID-19 cases among passengers of cruise ships where subsequent multi-jurisdictional outbreaks occurred. As at 31 May 2020, approximately one third of outbreak associated cases and one third of deaths were associated with cruise ships. These figures underestimate the actual number of cases associated with cruise ships due to incomplete reporting of outbreak data to COVID-Net during this period.

After initial epidemic control nationally, Victoria experienced a resurgence of COVID-19 from an outbreak associated with a quarantine facility [15]. Subsequent community transmission introduced SARS-CoV-2 into high-risk settings such as RACF, disability services, and health care. Outbreaks in education and childcare settings peaked when community transmission was highest during July 2020, suggesting that infections in these settings were likely driven by community transmission. Children less than 10 years old are thought to transmit SARS-CoV-2 less than adults and adolescents [16]. Australian and international evidence indicate transmission of the virus within schools is low and child-to-child transmission within the school setting is uncommon [17-20]. We did observe a high number of outbreaks in kindergarten, childcare and primary schools when compared to secondary educational settings, while outbreaks in secondary schools were generally larger in size. However, there were exceptions to these finding where a single outbreak in primary-secondary school was associated with 209 cases.

The high number of outbreaks in the workplace and industry settings during the second wave reflects the staging of lockdown measures. Essential workplaces that were not able to close services or transition workers home such as manufacturers, construction sites, and warehouses were disproportionately affected. Confined and shared spaces associated with a lack of physical distance and poor ventilation are known risk factors for outbreaks in workplace settings [21]. For example, workers in meat processing factories often socialise outside of work, share transport to work, and work in close proximity in processing lines in poorly ventilated areas [22, 23]. Although fewer outbreaks occurred in food industry settings, abattoir and meat processing plant outbreaks were large with a median of 88 cases (range 5–168) and 22 (range 3–212), respectively. The use of surveillance data to explore the role of superspreader events in settings where environmental conditions are conducive to SARS-CoV-2 transmission is limited. Better understanding of transmission in such workplace setting where infectious individuals work closely with many susceptible individuals, will help control the spread of SARS-CoV-2 as well as inform public health guidance [10, 11].

Transmission of SARS-CoV-2 among older people in RACF may result in significant morbidity and mortality due to age and a high prevalence of co-morbidities [24]. Early in the response, the Australian Government released national guidelines for COVID-19 outbreaks in RACF [25]. During the peak of the second wave, the number of outbreaks had surged to 88 outbreaks associated with 4,763 cases. During this time, facilities deployed major public health strategies to reduce the risk of entry and transmission of SARS-CoV-2 within facilities. Strategies include the development of outbreak management plans, additional infection control training of staff, and restricted visitor entry within facilities. Of RACF outbreaks across both epidemic waves, nearly half (46%, 397/853) involved 5 cases or less (Table 2). Nationally, outbreaks with longer duration were not necessarily those with the highest case numbers or deaths. This may be due to some facilities experiencing two outbreaks or having gaps of at least 14 days between cases.

## Limitations

These surveillance data are subject to several limitations. Firstly, NSW did not report data to COVID-Net for the entire surveillance period, and therefore the picture of outbreaks in Australia during the first wave is incomplete. Secondly, we cannot comment on outbreak risk by setting in the absence of denominator data. However, these data do provide insight into highly affected settings. Thirdly, there was potential for variation in the classification of where outbreaks occurred, although we attempted to standardise this through use of a common database with standardised definitions. In addition, the development of national guidance for screening and testing of cases and close contacts changed as understanding of SARS-CoV-2 transmission increased. Higher rates of screening of staff and residents in certain high-risk settings such as in hotel quarantine and RACF, inevitably improved outbreak detection in these settings resulting in a bias towards detecting cases and outbreaks. As surveillance data are subject to change due to reconciliation, outbreaks and case data reported in our report may vary from previously published reports. Finally, our definition of an outbreak requires two or more linked cases. States and territories have gradually moved to the presence of a single case in any setting as an outbreak definition [4].

## Conclusion

During 2020, Australia experienced fewer cases and outbreaks than many other higher income countries. This was the result of concerted efforts at all levels of society, including governments and industry, along with extremely high levels of community engagement. Efforts by jurisdictions to maintain robust isolation of cases and contact tracing capability was critical to prevent community transmission. Our findings demonstrate that multi-jurisdictional outbreaks of COVID-19 occurred regularly prior to domestic and international border closures and require a rapid and coordinated response. Strong public health policies on physical distancing and hygiene, particularly in high-risk settings, will assist in the prevention of outbreaks across all settings. As the rollout of COVID-19 vaccines occurs, it is vital outbreak investigation teams prepare to monitor changing transmission within Australia, particularly within closed cohorts of people. Robust outbreak investigations may indicate groups with lower vaccine coverage and will assist with estimating vaccine effectiveness. There are many lessons to learn from Australia’s experiences to prepare for and manage COVID-19 outbreaks in this period of high epidemic control. Ongoing surveillance and analysis of COVID-19 outbreaks will provide an important indicator in response to evolving policies and response strategies for Australia and other countries.

## Data Availability

All relevant data are within the manuscript and its Supporting Information files

## Acknowledgements

The COVID-Net surveillance network conducted surveillance of outbreaks in partnership with a range of stakeholders, particularly state and territory health departments. We thank the CDNA for their work to coordinate surveillance and response in Australia and review of this document. We thank the COVID-Net epidemiologists, project officers and interviewers at jurisdictional health departments who contributed to this report. We thank Rose Wright and Kate Ward from the Australian Government Department of Health and Tracey Tsang from NSW Health for their review of the manuscript. We acknowledge the work of various public health professionals and laboratory staff around Australia who tested specimens and investigated outbreaks. The quality of their work was the foundation of this report.

## Supporting information

**S1 Appendix. Outbreaks of COVID-19 by setting and sub-setting, Australia, 28 January – 27 December 2020**.

**S2 Appendix. COVID-Net definition of outbreak settings and sub-settings**.

